# A qualitative study exploring Emergency Department Staff attitudes to COVID-19 research

**DOI:** 10.1101/2024.07.04.24309781

**Authors:** Joanna Quinn, Antonia Ho, Andrew Blunsum, David J Lowe

## Abstract

**Introduction:** Research conducted in the Emergency Department (ED) is essential for improving patient care and advancing evidence-based practice. However, there are several challenges to research engagement in the ED, including lack of time, awareness of research opportunities, and concerns about the impact on clinical duties.

This study aimed to explore the attitudes and perceptions of ED staff involved in an enhanced syndromic surveillance of hospitalised severe acute respiratory illness (CHARISMA study) during the COVID-19 pandemic.

**Methods:** This qualitative study utilised semi-structured interviews with a mix of nursing and medical staff with a range of experience levels. Thematic analysis was then undertaken.

**Results:** 9 respondents informed our four key themes: the value of research, the user experience of the study tools, clinician research engagement and improvement recommendations for future iterations of the study. Our findings reveal that ED staff value research and recognise its importance in improving patient care and evidence-based practice. However, they also face significant challenges in participating in research due to time constraints, lack of awareness of research opportunities, and concerns about the impact on clinical duties.

**Conclusion:** To address these challenges, we propose strategies to enhance research engagement in the ED, including providing more support from senior staff, more transparent communication about research studies, training on research methods and tools, and opportunities for feedback and input. Implementing these measures, we can enhance the environment for research in the ED, enabling wider staff contribution.

**What is already known on this topic:** Data collected in the Emergency Department by clinical and research teams was an important contribution to COVID research including multinational platform studies. However, little is known about the attitudes of front-line staff involved in this process. This study responds to a paucity of literature with regards to clinician attitudes towards being asked to participate in research during the COVID-19 pandemic in an acute setting.

**What this study adds:** This study provides the first published description of ED staff attitudes to their involvement in research during a global pandemic. It highlights the challenges that are posed by the unique pressures of the ED setting - namely activity and unpredictability, time pressure, and in some cases, a lack of resources.

**How this study might affect research, practice or policy:** This study reinforces the need, value and support of staff to conduct research in an ED setting. We hope that this study provides useful insight to researchers on potential pitfalls to beware when undertaking their own original research. We anticipate that many of our findings are highly generalisable to other ED and acute care settings and hope that this encourages further expansion of this area.

## INTRODUCTION

The emergence of the novel coronavirus - SARS-CoV-2, posed significant challenges to healthcare systems worldwide[1]. As a society, we faced rapid adaptation to strict social distancing measures and significant impingements on our freedom of movement[2]. In secondary care settings, cohorting patients based on their probability of SARS-CoV-2 infection changed clinical practice, often making treatment decisions with a rapidly evolving evidence base [3]. Characterisation of the clinical features of SARS-CoV-2 (COVID-19) was required to support decision-making, develop clinical pathways and provide up-to-date information to clinicians as each variant emerged. Testing provision and disease management were initially limited, and there was an urgent need for real-time data to support and guide policy, resource allocation and clinical care pathways[4]. Previous respiratory surveillance had largely focussed on annual influenza outbreaks, and this provided the foundation for the initial response - with programmes of research and surveillance adapting to the pandemic and its unique challenges [5,6]. There was a clear need to integrate research into clinical practice with platform trials such as RECOVERY[7], providing critical evidence to support treatment decisions[8]. Worldwide efforts such as the International Severe Acute Respiratory and emerging Infection Consortium (ISARIC) Clinical Characterisation Protocol (CCP) study[9] supported harmonised data collection but were limited to research methodology, leading to the requirement for additional approaches to surveillance.

### Background

The Queen Elizabeth University Hospital (QEUH), Glasgow, is a tertiary adult (>16 yrs) hospital with approximately 110,000 emergency department attendances per year with specialist infectious disease service onsite. During the initial phases of the pandemic, QEUH had some of the highest admission rates in Scotland as well as the highest mortality rate [10]. In response to the need for surveillance - the CHARISMA study[11] was initiated. The study was a prospective, observational cohort study, utilising routinely collected data alongside the creation of an electronic patient record (EPR). This provided near real-time data reporting on hospitalised severe acute respiratory illness (SARI) to Public Health Scotland (PHS) and weekly updates to frontline staff. Informed consent for participation was not required as the clinical data that was collected and the respiratory viral PCR testing was that of standard care. Appropriate ethical approval was also sought prior to commencement of data collection.

A specially developed ‘SARI Turas Clinical Assessment Tool’ was developed by NHS Education for Scotland (NES) and was integrated into the EPR. Clinical staff were trained in its use and supported by the ED research team. Patient demographics, presenting symptoms, COVID-19 & flu vaccination history, patient journey, and outcome were recorded. An ‘order set’ was created within Trakcare (the EPR) to allow standardisation of test requests by clinicians. All patients underwent point-of-care testing for influenza A/B & SARS-CoV-2 PCR (cobas® Liat®). Linkage to routine healthcare data provided supplemental information on co-morbidities, medication history, and laboratory and radiology results. A dashboard (Microsoft Power BI, Washington, USA) with patient demographics and weekly influenza A/B & SARSCoV-2 PCR positivity was available to Public Health Scotland (PHS) and hospital clinicians in real time. [11]

During the study period of CHARISMA, it was challenging to ensure that consistent recording and testing was performed by clinical staff. The Turas Clinical Assessment Tool (TCAT), intended to be completed by the admitting clinician was variably completed at the time of assessment, with clinicians reverting to paper notes. This increased the workload for the research team, who were tasked with retrospectively completing the missing data sets alongside the other elements of running the study.

In anticipation of future research and surveillance, for SARS-CoV-2 and future, novel viruses or outbreaks, this study aimed to explore the attitudes and perceptions (including perceived facilitators and barriers) to engagement with the CHARISMA study amongst clinical staff working in the Emergency Department.

## METHODS

This was a qualitative study involving semi-structured interviews with staff members currently working in the Emergency Department (ED) at QEUH, Glasgow who participated in the CHARISMA trial during its initial study period (Nov 2021 - May 2022). All participants were aged over 18 years. Data collection occurred during June 2022 and July 2022.

### Recruitment

Study participants were recruited following an email invitation to staff working in the ED from various staff groups, including junior doctors, consultants, research nurses and staff nurses.

The sample is a convenience sample of those who responded to our request. We used purposive sampling to recruit a variety of staff members from different clinical backgrounds.

### Data collection

Interviews were conducted by one researcher (JQ) either face-to-face or online using Microsoft Teams software. All interviews were conducted one-to-one, with no one else present. Most participants were interviewed in the workplace. A semi-structured topic guide was developed by the study team (JQ and DJL) to guide the discussion – this was not pilot tested prior to study commencement. The guide focused on two elements: i) attitudes towards research studies in the Emergency Department during the COVID-19 pandemic; and ii) the use of the specially designed tool integrated into clinical practice to support data capture. (see Supplementary material)

Participant information sheets and consent forms were given to those wishing to participate before data collection. Verbal consent was also obtained at the beginning of each interview.

The interviews were recorded with participant consent, with professional transcription then performed to anonymise the outputs. A transcript of each interview was made available to the research team however transcripts were not given to participants and no repeat interviews were undertaken. Interviews were conducted until the research team felt they had reached the saturation point of new ideas/themes.

No participants chose to withdraw their involvement following their interview participation.

### Data Analysis

Analysis followed the framework approach: team members listened to the interviews and read the transcribed interviews to gain familiarity with the content. A thematic framework was developed iteratively throughout the interviews by JQ, with DJL reviewing additional transcripts. An inductive approach was taken with emerging themes identified. The thematic framework was then applied to each interview by JQ. Findings were shared with the research team, with a discussion of interpretation.

### Patient and Public Involvement

As this study focused solely on the attitudes of Emergency Department staff members and the research team, there was no patient and public involvement.

## RESULTS

### Respondents

We recruited 9 key participants to inform our study over a period of 2 months. We aimed for a broad spread of backgrounds, including nursing and medical staff. (Table 1) The interviews lasted between 7 and 17 minutes.

**Table 1.** Characteristics of participants.

| Characteristic | Number of participants |
| --- | --- |
| Gender<br>• Male<br>• Female | 2<br>7 |
| Age (years)<br>• <30<br>• 30-60<br>• >60 | 4<br>5<br>0 |
| Role<br>• Nurse<br>• Junior Doctor<br>• Research nurse | 3<br>4<br>2 |
| Interview<br>• Face to face<br>• Online | 6<br>3 |

**Table 2.** Table of themes and sub-themes.

| Value of research | User experience of research tools | Research engagement | Suggestions for future iterations |
| --- | --- | --- | --- |
| Patient care | Knowledge | Awareness | Knowledge |
| Staff participation | Process | Value / perceived benefits of the trial | Process |
|  | Testing | Enablers to engagement | Testing |
|  |  | Barriers to engagement |  |

### Value of research

All respondents felt that research conducted in the ED was of value, both in terms of improving patient care as well as contributing to evidence-based practice. There was, however, concern that it should not take staff away from their clinical duties and that embedding research into routine practice requires resourcing, increased staff levels and departmental motivation. There was agreement that ED provided an excellent opportunity to identify (and potentially recruit) a wide variety of patients at the very beginning of their admission, and that this was perhaps underutilised currently.

> *Respondent 3: “Research is a thing that we can do to change practice, to ensure clinical excellence, and actually change pathways in a very pragmatic way and all for the better.”*

> *Respondent 6: I” think …. we all have a duty as doctors to help out with research because it’s how we improve care for our patients.”*

Intrinsically linked to the value of research is the theme of staff engagement; respondent 9 suggested, “*it is important, and I think making people aware that it’s something that happens continually is important as well. Like our improvement is for everyone. It’s not just for specific people”*.

### Clinician experiences

Exploring the clinician experience of participation in the CHARISMA study identified three sub themes:

- Knowledge (awareness of study, onboarding to trial, etc)
- Processes (data input, use of TURAS tool, patient enrollment)
- Testing (use of swabs and samples)

### Knowledge

There was significant variation between respondents about the degree of awareness they had about the study and the level of preparedness they felt they had. This was despite regular efforts being made to ensure all staff were informed, including development of a clinical pathway and Standard operating Procedure (SOP) handbook, regular reminders and prompts at handover and induction for new staff.

> *Respondent 9: “so I think for a while we were really focused on recruiting for CHARISMA. It was often brought up at handovers and just, like, gentle reminders, like everyone was pretty good at it for, you know, a couple of weeks here and there and then it would kind of tail off and the reminders would come back and then we would back again, being a bit more focused on it*.

> *Respondent 7: (regards recruitment) “The only information I really had was that they were trying to recruit anyone with a, sort of, acute respiratory illness. I didn’t really, I don’t think I had any talk at the start of it as to what exactly, whether there were more inclusion criteria than that but essentially, that was the message that I felt was reinforced was that anyone with an acute respiratory illness should be put through as a possible CHARISMA patient.”*

This perhaps reflects the difficulties surrounding information dissemination in a 24-hour, shift-based service. However, respondent 6 reported, “*it was probably the one that I had the most awareness of [out of the multiple studies running in the department concurrently].”*

These responses demonstrate the variance in the knowledge of staff members, both around the study - CHARISMA - as well as around research more generally in the department. Where multiple studies and trials are running concurrently, there is a risk of information overload or saturation, often competing interests for attention, compounded by the time pressures so frequently experienced recently.

Respondent 3 and 4 both reflected on how to maintain awareness:

> *Respondent 3: “We’ve had a huge turnover in staff …., and I think having research as part of …. induction is of the utmost importance….”*

> *Respondent 4: “I think you need the kind of support from your seniors on shift as well and to be aware of flagging patients that it can take extra time to do the things that are necessary.”*

### Processes

The Clinical Assessment Tool was designed to follow clinical workflow from patient demographics, background, COVID-specific features and assessment. Feedback regarding the CAT reported it was straightforward and easy to use. It was noted, however, that there was some duplication of documentation and an increase of the ‘administrative burden’.

> *Respondent 1: “I found it very straightforward, and [it] didn’t take up much time….. The rest of the details were pretty much in the top of your head anyway because you were seeing the patient, so it was quite easy, and … didn’t require any patient consent …. so it was quite straightforward.”*

Concerning patient enrolment, it was felt to be easy to recruit to due to the broad inclusion criteria. Staff were also often supported by the research team to recruit/enrol patients. Senior support on the shop floor was cited as a facilitator or prompt for enrolment however sometimes the opposite could be said if the consultant in charge was less research aware.

> Respondent 6: *“I think the team…the senior decision-makers who are aware of what research is going on in the department obviously had no problem with it at all, and in fact would say, actually, I think this patient would be eligible for this, and then tell you how to go and recruit them for that. But the ones who were less aware of what’s going on, it was just…it didn’t really cross their mind.”*

> *Respondent 9: “It wasn’t difficult to recruit to CHARISMA. I think there was a couple of extra added steps … but most of the time it was easy enough to just include it in your kind of routine set-up and also the patients.”*

Once staff were familiar with the process for enrolment, the clinical assessment tool was found to be straightforward and user-friendly. The development team were keen that this should not be a barrier to recruitment and enrolment and were careful in the design of this tool. There was also rigorous user testing prior to the study commencing.

### Testing

Regarding testing, there was a range of responses reflecting responders’ differing job roles alongside the evolving testing requirements. A key theme identified was around the complexity of the tests required and the confusion surrounding the use of point-of-care testing. During the study period, there was also changing guidance concerning indications to perform the test. An integrated order set within the EPR was, however found to be helpful.

> *Respondent 4: “I also think one of the things … was when patients were being front-loaded for bloods and things, whether that was getting through to the nursing staff and the HCA’s [*health care assistant] *that they needed to have the CHARISMA set done. There was confusion over where the point of care should be done on people when we weren’t normally doing point of care.”*

> *Respondent 1: “So I think that’s confused some people because the standard of care changed during that study to all the patients that were coming in with some sort of respiratory illness were getting the COVID swabs. So I think that probably stopped some people getting the COVID swab that you might have wanted to get the swab.”*

### Engagement with the CHARISMA Study

Within the topic of research engagement, we identified themes around four main topics:

- Awareness
- Value / perceived benefits of the trial
- Enablers to engagement
- Barriers to engagement

### Awareness

Participants reported that there was good awareness of the study with visual cues (posters) as well as reminders on the shop floor and at handover. There was agreement that sometimes, with multiple studies running simultaneously, there can be an overload of information, and there were perhaps periods where reminders and awareness ‘tailed off’. The research nurses were highlighted as maintaining awareness of and supporting recruitment to the study.

> *Respondent 2 :”I think it’s just having an awareness of it and I think it probably just needs to be consistently highlighted. I think doing it once or even twice is not really enough because people forget about it, and if people are on leave or on different shifts, they don’t always read their emails. I think having the single poster with all the department’s research studies on it has been quite useful, because it’s got quite easy to access information on it…“*

### Perceived benefits

Having the results of the trial disseminated to staff was felt to be a positive reinforcement of the value of participating in research - respondent one states …” *it shows people why you’re doing [it] and it’s quite interesting and I think useful to engage people.”*

> *Respondent 3: “I actually think that needs to begin at university level … The nurses’ understanding of research is reading qualitative versus quantitative papers and trying to compare and contrast …. but it’s not what true research is about. They don’t get a true understanding of how important clinical research is, and how it impacts how we perform as nurses. And promoting these positive patient stories, I think, will then give nursing staff incentive and a boost to also think, oh yeah, there’s a trial to do with that.”*

> *Respondent 5: “No, I think the research things are important, but then I think undercutting that has to be, do the staff see the benefit of it? And I think with these studies being quite long, and I’m not aware we always see the initial benefit, it was quite short term, in that way. There are longer-term studies required, but I don’t think we always see the short-term benefit of it, and that’s for research fellows to argue about, down the line. But we don’t see the short-term benefit of it. I think, if you were made aware of what has been changed because of, then maybe it will encourage you more to be more active in that.”*

### Enablers to engagement

All respondents felt that the research nurses were a huge help during the study period. They were able to support team members to recruit patients to the study as well as identify eligible patients. There was, however a suggestion that their presence was missed during out-of-hours working, perhaps when, actually, more support is needed.

> *Respondent 4: “I think what is good in this department is you have the dedicated research nurses. They are always there to flag and recognise patients coming through the door, what is it, 8-5 when they work.”*

> *Respondent 2: “I think having the research nurses in the department was quite useful because I at one point asked one of them, and especially when they’re kind of circulating around the department or if they come and identify a patient that you’ve seen, I think that’s very useful because they, kind of, take you through the process.”*

The concept of incentives was also mentioned, including a certificate of involvement or allocating roles, e.g. ‘CHARISMA champion’.

> *Respondent 2: “I think probably the other thing with any of the research studies is that it’s useful for the more junior medical staff to get involved in as well, and I think the kind of, way of providing some sort of evidence of participation is useful for them.”*

### Barriers to engagement

Discussion around perceived barriers to engagement included busyness of the department (mentioned by five respondents), reduced staffing levels (particularly at night) and time taken to perform the additional tasks required for the project.

> *Respondent 5: “Night variation: I think massively, but then there’s also the variation of staffing from the department. So, once we became aware of the SARI order set, what patients were generally requested in that group for it, then we would try to do it through the night, but there will be an obvious variation in nights and days.”*

> *Respondent 8: “But I think during the week, during the day, it always seemed a lot more doable than… out of hours.”*

Respondent 2 cited the duplication of efforts if they had already had investigations but required further samples to be sent as a barrier to engagement:

> “*I think time is a bit of a factor, like particularly if it was extra bloods or swabs and things that needed taken. Like if the patient had already been bled and then they might have needed something extra sent. That is a time factor of who’s going to do that, because I didn’t always have time and the nurses didn’t always have time either, so then it was not always done. I think there were probably some that were missed out because of that.”*

A recurring theme was being able to remember to recruit to the study alongside ensuring that all staff members had a good understanding of the study and recruitment criteria.

> *Respondent 9: “You definitely did find variation depending on who was on. Whether that was a nurse in charge or whether that was a consultant in charge.”*

> *Respondent 1: “I think the busyness of the department, you get quite focussed clinically when you’re doing clinical stuff and all, kind of, research goes to the back of your mind unless it’s your own research” [Editor note - even if it is your own research!]*

### Suggestion for future iterations

Many respondents had suggestions regarding how to improve future integrated surveillance studies. These were, again, grouped into themes around knowledge, processes and testing.

### Knowledge & awareness

- Increased number of research nurses with extended hours
- Engagement of all staff members, including healthcare support workers and nursing staff
- Daily reminders in handover and at team briefs
- Newsletter/weekly brief information

> *Respondent 5 “I think it has to be multifaceted. I think there has to be social media communication, there has to be postering, there has to be the word-of-mouth thing, certainly, but postering, social media, and just an insistent drive to keep mentioning it, keep pushing it, that then improves the staff pick-up of it.”*

> *Respondent 3: “I think the newsletter, that unfortunately we haven’t had out the last few months, should have a big research part in it, and should make it more sexy, more appealing.”*

> Respondent 4: “*I think it’s just about putting it at the forefront of the whole team’s mind isnt it?”*

### Processes

- Suggestion of pro-forma to reduce duplication of efforts.
- Reducing the cognitive load for clinicians!

> *Respondent 1: “Maybe a respiratory illness pro-forma”*

> *Respondent 4: “Other than like electronic alerts on TrakCare and things, but then you already have quite a lot of stuff on TrakCare already.”^1^*

> *Respondent 1: “I think relying on clinicians as little as possibly is probably the best way forward.”*

### Testing

- Streamlining order sets and increasing visibility on Trakcare
- Simplifying ordering of microbiology ordering (often required extra information when requesting)
- Ensuring triage were able to identify patients from the outset to ensure correct tests ordered (perhaps tool or guideline

> *Respondent 2: “I think in terms of research as well it doesn’t always have to come from the medical staff. Like it would actually be really useful if the nursing staff highlighted people early as well or asked do you think they’re suitable?”*

> *Respondent 6: “It would be good if there was a way to kind of set up the cultures and things so that it’s already pre-populated with the right information, but I don’t know if TRAC allows for that. I: I know, and things like the swabs, having to manually put in like nasopharyngeal or throat or…yeah. R: Yeah, and having to remember that one of the…you know, you’ve got a blood culture and a sputum culture and stuff as well.”*

## DISCUSSION

Multiple themes were identified during the data analysis relating to both delivery of the CHARISMA study as well as some related to more general attitudes towards research. Our results demonstrated a high level of agreement amongst participants in terms of the positive value of conducting research in the Emergency Department; however, they also described themes associated with the practicalities of performing research in an ED setting. These findings must be set within the context of this study being conducted during and related to the COVID-19 pandemic and the unique circumstances that this brought. Medical research gained greater prominence both within clinical areas but also the wider public due to vaccine research and platform studies such as RECOVERY to identify optimal treatment and epidemiological reporting ^7^. The most important challenges faced were cited as the difficulty of maintaining staff awareness and engagement. This was often multifactorial but included the high turnover of staff in the department due to the rotational nature of junior doctor’s working patterns, the shift-based, 24 hour service, alongside departmental factors such as crowding and lack of flow. It was also felt to add extra time (either perceived or actual) to complete the necessary paperwork or laboratory samples and staff felt that it was increasing their workload somewhat.

Whilst ED crowding is now a common occurrence, the busier the department was, the greater the impact this appeared to have on patient identification, reflecting the stressful and busy environment we work in and perhaps the unpredictable nature of our work at times. Often, things which are perceived to be ‘added extras’ and not directly related to patient care (such as research) can be deprioritised in this context.

The presence of research nurses within the department was felt to be extremely positive and a key enabler to successful patient participation. This team provided support, guidance and expertise that allowed the study to continue, despite some of the challenges faced.

Of those clinicians who did recruit patients to CHARISMA, and utilised the TCAT tool, there was agreement that the inclusion criteria and enrolment were straightforward, and that the tool was simple to use. This highlights the importance of utilising or developing tools and software that enhance user experience and fit within staff members current clinical workflow. Of note, the changing sampling guidance was cited as confusing and unclear however was outside the control of the study team and was, in part, influenced by national availability of POC tests.

Considering the future possibilities of syndromic surveillance in the ED, it is very much possible. However, it will take concerted effort, with widespread staff training and engagement. Clear instructions and guidance need to be provided, with a consistent message from the research team and senior staff members to ensure recruitment and participation occur, despite how busy or crowded the department might be. Incentives to maintain engagement may be as simple as sharing the interim findings with staff regularly, as well as providing important evidence for portfolios.

### Comparison with previous literature

There is paucity of research available exploring the attitudes of Emergency Department staff towards the process of research. A review article by Johnston et al[12] makes reference to the stressful and busy nature of the Emergency Department environment and commented on the unpredictable nature of the job, which is reflected in our findings, however did not expand further to include their opinion on research. A survey of EM trainees in Australia from 2017[13] did touch upon barriers to performing research, however this was neither site specific, nor study specific and focussed solely on EM trainees across a broad region. Despite this, the main barriers identified included time and skills, but also a lack of research mentoring, training and research positive culture. These reflect some of our study’s key findings.

A piece of work from the clinical research nurses in Edinburgh[14] based at the Royal Infirmary of Edinburgh and part of the Emergency Medicine Research Group Edinburgh (EMERGE) highlighted some of the challenges their research nurses experienced whilst conducting research in an ED setting, these included factors such as interruptions to tasks, competing interests such as diagnostic or therapeutic interventions and patient variability, all of which were prevalent in our study findings.

This study has identified both possible barriers and incentives to consider whilst undertaking research in an Emergency Department setting. It supports the view that we *should* be doing research in the ED and that in general, most staff are supportive of this. We hope our findings support future researchers to consider optimal approaches for delivery. We would urge participation in the development of strategies to engage and encourage staff, ensuring all staff members are informed and included – especially during times of rotational changes. A particular consideration within the ED is developing means to update staff on the progress of the study and to inform them of any protocol changes or developments that they need to be aware of. The presence of research nurses in the department was cited as positive and helpful by many participants and should be considered where possible. Recognition of their positive impact on study recruitment and staff support should be balanced by their hours of work, with poorer performance and often busier departments overnight meaning recruitment is limited to, often, daytime.

### Limitations

This is a single site study with 9 participants therefore findings should be interpreted within this limitation. This work provides a snapshot of staff members’ views and experiences at that time, but should be taken in the context of research being conducted during a global pandemic - a time of unprecedented stress on acute care services. We recognise that their insights and opinions may have been significantly influenced by the confounding factor of COVID-19 - with all the extra challenges that this presented. Convenience sampling allowed us to include a range of staff from varying backgrounds and with different experiences however there is always the risk that other staff members did not get a chance to share their (perhaps differing) views.

We also recognise the risk of selection bias amongst the participants, as well as the potential for recall bias (as interviews were conducted some months following the end of the study). Finally, the researcher (JQ) being a fellow staff member and colleague may have unknown positive and negative effects on participants’ honesty and openness.

Despite these limitations, we are optimistic that our findings remain valid and relevant, and reflect the mood and feeling of the time.

## Conclusion

This study highlights some of the challenges faced when trying to conduct research within an Emergency Department setting. Although our study was specifically related to the implementation of the CHARISMA study during the study pandemic, we believe that it is generalisable to many other instances of data collection in an Emergency Department setting. It touches upon the busy and unpredictable nature of the ED workflow as well as considering the 24-hour rotation of staff and the difficulty of dissemination of information. It highlights a consideration about ‘whose role is it to perform research anyway?’ and raises the question of not only how to engage a more significant proportion of staff (especially non-medical staff) in research alongside showing them the benefits of involvement.

It reiterates the need for a supportive and research-positive environment as well as the need for a dedicated research team to support staff with high visibility and strong leadership. Promoting a culture where research is not only routine practice but is executed to a high standard should be something that all departments strive for for the benefit of our patients.

## Supporting information

COREQ Checklist

## Data Availability

All data produced in the present study are available upon reasonable request to the authors

## Author Contributions

AH / DJL / JQ / AB conceived the study. JQ led analysis supported by DJL. JQ wrote the first draft. JQ, AH and DJL edited and approved the final manuscript.

## Acknowledgements

The authors would like to thank all of the colleagues who agreed to be involved in this study. We would also like to thank the research nurses at the QEUH, without whom, nothing would be achieved.

## Data availability statement

Data is available upon reasonable request.

## Ethics statement

Ethics approval for this sub-study was included within the ethics approval for the CHARISMA study: obtained from the North of Scotland Research Ethics Committee (ref. 21/NS/1036) and the NHSGGC Biorepository (ref. 16/WS/0207) to store residual blood and respiratory samples. The participants gave informed consent to participate in the study.

## REFERENCES

1. WHO. WHO director generals opening remarks at media briefing on COVID-19. https://www.who.int/director-general/speeches/detail/who-director-general-s-opening-remarks-at-the-media-briefing-on-covid-19---11-march-2020.

2. UK Government. Staying at home and away from others. https://www.gov.uk/government/publications/full-guidance-on-staying-at-home-and-away-from-others. Updated 2020. Accessed 21/11/, 2023.

3. UK Government. Coronavirus action plan 03.03.2020. https://www.gov.uk/government/publications/coronavirus-action-plan/coronavirus-action-plan-a-guide-to-what-you-can-expect-across-the-uk.

4. UK Health Security Agency. Coronavirus (COVID-19): Using data to track the virus. https://ukhsa.blog.gov.uk/2020/04/23/coronavirus-covid-19-using-data-to-track-the-virus/. Accessed 21/11/, 2023.

5. McLeish NJ, Simmonds P, Robertson C, et al. Sero-prevalence and incidence of A/H1N1 2009 influenza infection in scotland in winter 2009–2010. PLOS ONE. 2011;6(6):e20358. 10.1371/journal.pone.0020358.

6. Tolksdorf K, Buda S, Schuler E, Wieler LH, Haas W. Influenza-associated pneumonia as reference to assess seriousness of coronavirus disease (COVID-19). Eurosurveillance. 2020;25(11):2000258. https://www.eurosurveillance.org/content/10.2807/1560-7917.ES.2020.25.11.2000258. doi: 10.2807/1560-7917.ES.2020.25.11.2000258.

7. Wilkinson E. RECOVERY trial: The UK covid-19 study resetting expectations for clinical-trials. BMJ. 2020. http://www.bmj.com/content/369/bmj.m1626.abstract. doi: 10.1135/bmj.m1626.

8. Scottish Government. Coronavirus (COVID-19) phase 1: Scotland’s route map update. https://www.gov.scot/publications/coronavirus-covid-19-framework-decision-making-scotlands-route-map-through-out-crisis-phase-1-update/pages/2/.

9. ISARIC - COVID-19 clinical research resources. https://isaric.org/research/covid-19-clinical-research-resources/ Web site.

10. BBC. Covid in scotland: QEUH records the most covid deaths… Available from: https://www.bbc.co.uk/news/uk-scotland-57256232.

11. Ho A, Blunsum A, Quinn J, et al. A novel syndromic surveillance of influenza and SARS-CoV-2 infections among patients with hospitalised severe acute respiratory illness in glasgow, scotland, november 2021 to may 2022. ISIRV Conference Abstract. 2022.

12. Johnston A, Abraham L, Greenslade J, et al. Review article: Staff perception of the emergency department working environment: Integrative review of the literature. Emergency Medicine Australasia. 2016;28(1):7–26. 10.1111/1742-6723.12522. doi: 10.1111/1742-6723.12522.

13. Olaussen A, Jennings PA, O’Reilly G, Mitra B, Cameron PA. Barriers to conducting research: A survey of trainees in emergency medicine. Emergency Medicine Australasia. 2017;29(2):204–209. 10.1111/1742-6723.12734. doi: 10.1111/1742-6723.12734.

14. O’Brien R, Black P. Researching participant recruitment times. Emerg Nurse. 2015;23(7):26, 28-30. doi: 10.7748/en.23.7.26.s27.

